# Quantitative, Epitope-specific, Serological Screening of COVID-19 Patients Using a Novel Multiplexed Array-based Immunoassay Platform

**DOI:** 10.1101/2020.09.25.20201269

**Authors:** JM Blackburn, ND Anuar, TM Tan, AJM Nel, M Smith, K Ellan, NIS Bahrin, NSM Rosli, NH Badri, TNA Rahman, A Anwar, RM Zain

## Abstract

Following the COVID-19 pandemic outbreak in late 2019, a large number of antibody tests were developed for use in seroprevalence studies aimed at determining the extent of current or previous SARS-CoV-2 virus infections in a given population. The vast majority of these tests are qualitative and use a single target for antibody detection, incorporating either full-length or truncated versions of the nucleocapsid (N) or spike (S) proteins from SARS-CoV-2. Importantly, mono-epitope tests – whether qualitative or quantitative - are unable to localise antibody binding or characterise the distribution and titres of epitope recognition by anti-SARS-CoV-2 antibodies within an individual or across a population. However, it seems plausible that if such information were available, it may correlate with the presence of potent, high-titre, neutralising antibodies that afford protection again imminent re-infection, as well as with the likelihood of developing a memory B-cell response that would provide more durable protection. We have developed a novel, quantitative, multi-antigen, multiplexed, array-based immunoassay platform, ‘ImmuSAFE COVID+’ (ImmuSAFE) comprising 6 functionally validated domains or regions of the N protein of SARS-CoV-2 expressed using Sengenics’ KREX technology. This array platform enables determination of both the position and breadth of anti-SARS-CoV-2 antibody responses following natural infection or vaccination. To validate our platform, 100 serum samples (confirmed sero-positive COVID-19 cases, n=50; pre-pandemic HIV positive controls, n=50) were tested for IgG seropositivity to the N antigen, yielding 100% specificity and 100% sensitivity. All 50 cases showed positive antibody reactivity towards at least one N protein epitope, whilst all 50 controls showed antibody reactivity below threshold values. Broad variation was also observed in the magnitude and breadth of antibodies present, represented as an Epitope Coverage score (EPC). A positive correlation was observed between increasing age and EPC values, with individuals under 40 years old having a mean EPC score of 3.1, whilst individuals above the age of 60 had a mean EPC of 5.1. This finding may have broad implications for the natural history of COVID-19 disease in different individuals.

---

There are currently over 400 commercially available immunoassay-based tests for COVID-19 (Foundation for Innovative New Diagnostics (FIND), 2020), with rapid antibody and ELISA based tests constituting the majority. In most cases, these tests utilise a single antigen (mono-epitope) approach to determine qualitatively the presence of antibodies against SARS-CoV-2 viral antigens. Potential problems with false positives due to cross-reactivity between SARS-CoV-2 antigens and anti-SARS-CoV-1 antibodies, as well as antibodies against other common human coronaviruses, have been previously reported (Ma, Li, Ji, Ikram, & Pan, 2020; To et al., 2020; van der Heide, 2020). Given the heterogeneity of antibody responses within each individual towards different epitopes and antigens of SARS-CoV-2; (Gray, Peter, Mendelson, Madhi, & Blackburn, 2020) re-engineering and testing of functional epitopes of SARS-CoV-2 antigens has previously been utilised as a strategy to reduce false positives, but risks decreasing sensitivity through inadvertent engineering out of true epitopes in the targets. In addition, discontinuous epitopes in target antigens are often disrupted on absorption on to plastic or nitrocellulose surfaces, further reducing the available repertoire of biologically-relevant epitopes for antibody binding.

Screening for immunoreactivity in a high-throughput antigen microarray format potentially solves these problems by enabling multiple discrete, folded domains and epitopes of given target antigens to be assed simultaneously in a highly multiplexed, fully quantitative format that preserves the 3D structure of the antigen through the assay. Such a platform should facilitate the high throughput screening of COVID-19 samples for diagnosis of current or prior infection, as well as for quantitation and localisation of antibody binding, thereby providing a means to identify antibody correlates of on-going protection and development of durable immunity against future SARS-CoV-2 infection.

Viral proteins represent immune targets for both cellular and humoral responses and can thus be utilised in various biotechnology applications, including diagnostic assay development, immunosurveillance and vaccine development (Tay, Poh, Rénia, MacAry, & Ng, 2020). The use of the SARS-CoV-2 N protein in most COVID-19 immunoassay kits, for example, offers the ability to distinguish between COVID-19 patients and healthy controls due its antigenicity and immunogenicity (Hoffmann et al., 2020; Hofmann et al., 2004; Hofmann & Pöhlmann, 2004; Lin et al., 2003; Schoeman & Fielding, 2019). In addition, specific B-cell epitopes on viral surface proteins can also be used to detect protective/neutralising antibodies against SARS-CoV-2 (Ahn et al., 2020; Arabi et al., 2016; Shen et al., 2020; Sheridan, 2020).

The Sengenics ImmuSAFE COVID+ protein microarray was developed as a high-throughput immunoassay containing 6 functionally validated SARS-CoV-2 N protein domains (N core domain, N-terminal domain (NTD), C-terminal domain (CTD), intrinsically disordered region epitope 6 (IDR) epitope 6), intrinsically disordered region epitope 8 (IDR epitope 8) and intrinsically disordered region epitope 16 (IDR epitope 16)). This array leverages the KREX protein folding technology, which utilises an in-frame Biotin Carboxyl Carrier Protein (BCCP) as a folding marker and oriented immobilisation tag for recombinant antigens of interest. Full-length and functional domain proteins are expressed as fusions to the BCCP folding marker which becomes biotinylated *in vivo* by host biotin ligases only when the protein is correctly folded. Conversely, misfolded proteins drive the co-translational misfolding of BCCP, thereby blocking *in vivo* enzymatic biotinylation. This binary pattern of biotinylation ensures that only correctly folded, therefore functional, proteins are attached to a streptavidin-coated hydrogel support. (Adeola, Smith, Kaestner, Blackburn, & Zerbini, 2016; Blackburn & Shoko, 2011; Duarte & Blackburn, 2017).

In the present study, our objective was to quantify antibody titres, as well as to determine the position and number of discrete antibody epitopes recognised on the SARS-CoV-2 N protein in confirmed COVID-19 cases. We also aimed to determine the sensitivity and specificity of the ImmuSAFE COVID+ test through analysis of 100 serum samples provided by the Institute for Medical Research (IMR), Ministry of Health, Malaysia. Sera from fifty positive samples (cases) were collected from warded COVID-19 patients from Hospital Sungai Buloh, Selangor, Malaysia, as part of standard of care. The age of the patients in the cohort is summarised in Table 1. Fifty pre-pandemic HIV positive serum samples were used as true negative controls. All samples were assayed on ImmuSAFE arrays at a 1:50 serum dilution in the presence of 1% milk powder. Bound IgGs were detected and quantified as described previously (Lewis et al., 2018).

**Table 1.** Sample distribution by age and gender

| Age | Total | M | F |
| --- | --- | --- | --- |
| 20-40 | 10 | 3 | 7 |
| 41-60 | 24 | 21 | 3 |
| 61-73 | 9 | 6 | 3 |
| Unknown | 7 | 7 | 0 |

Figure 1 shows a strong and significant increase in antibody titres towards the N protein domains in the case samples relative to controls. The antibody concentration in serum was determined as a reciprocal titre, as described in the methodology section (Supplementary Materials, bioinformatics analysis). Reciprocal titres were scaled within a range of 0-100 (the highest reciprocal titre value being 57058.7 and 53412.3 for the N proteins and IDR regions respectively). The breadth of immune response in individuals was determined by the sum of the number of domains and epitopes which are positive for antibody reactivity, as represented in Figure 2. The dot plot in Figure 2A shows the diversity of the antibody response exhibited by different COVID-19 patients. In this analysis, an individual was considered to have positive IgG towards SARS-CoV-2 if the serum antibodies bind to at least 1 antigen on the array with a signal above the threshold determined by statistical analysis of the control sample data. Importantly, the data from the ImmuSAFE arrays confirmed that all 50 cases had quantifiable anti-N antibody titres, whilst none of the pre-pandemic controls had quantifiable anti-N antibody titres, equating to a sensitivity of 100% for known seropositive COVID-19 samples and a specificity of 100%.

**Figure 1:**
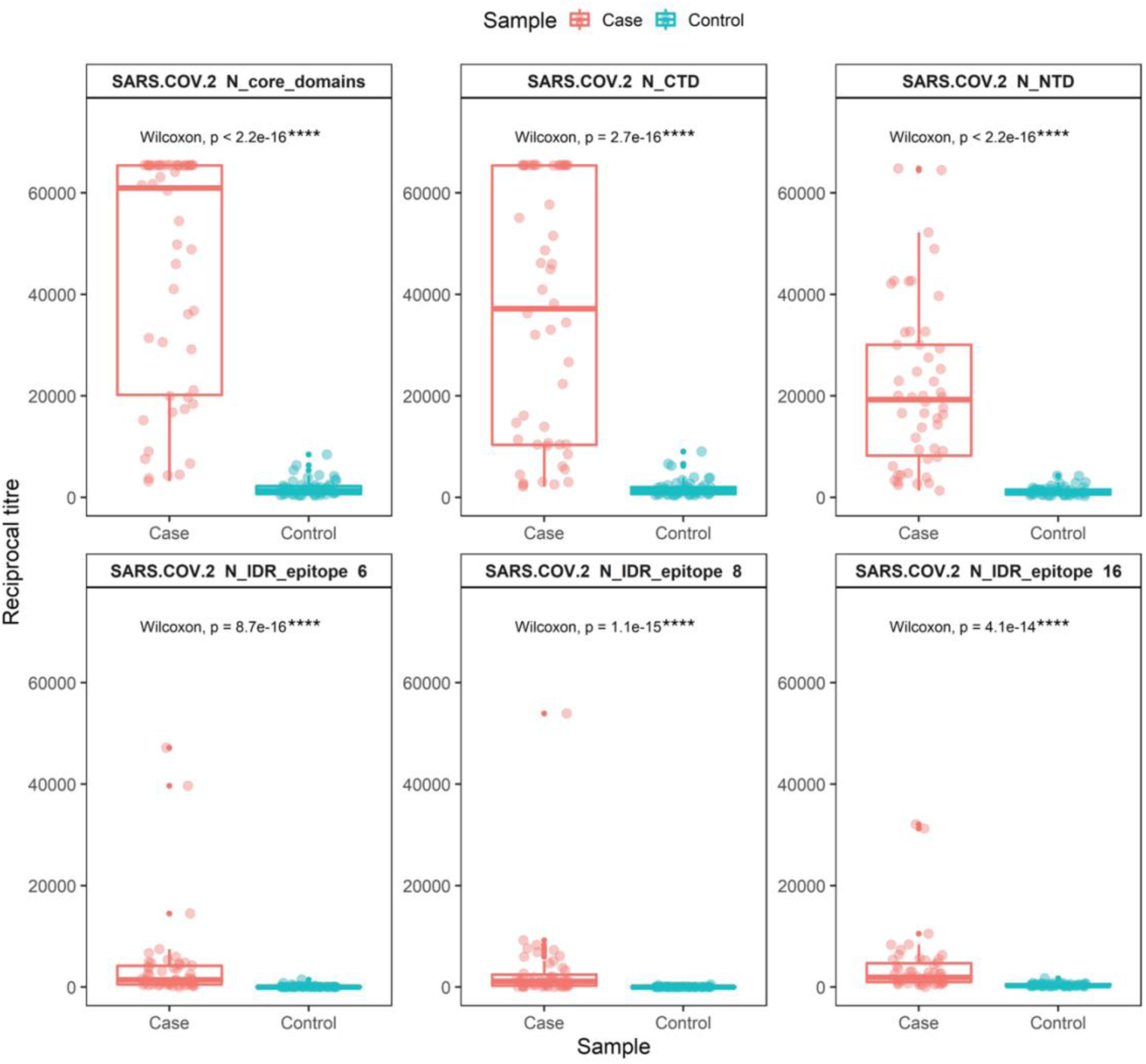
Overall distribution of antibody titre of samples tested (n=100) in each domain of the N protein (total domain, n=6). The sum of the reciprocal was used to determine the overall antibody titre against SARS-CoV-2 N protein epitopes and domains. Statistical analysis showed a significant (p<0.05) elevation of antibody titre in positive samples (case). P-values were determined using the Wilcoxon test method (unpaired, two-tailed).

**Figure 2:**
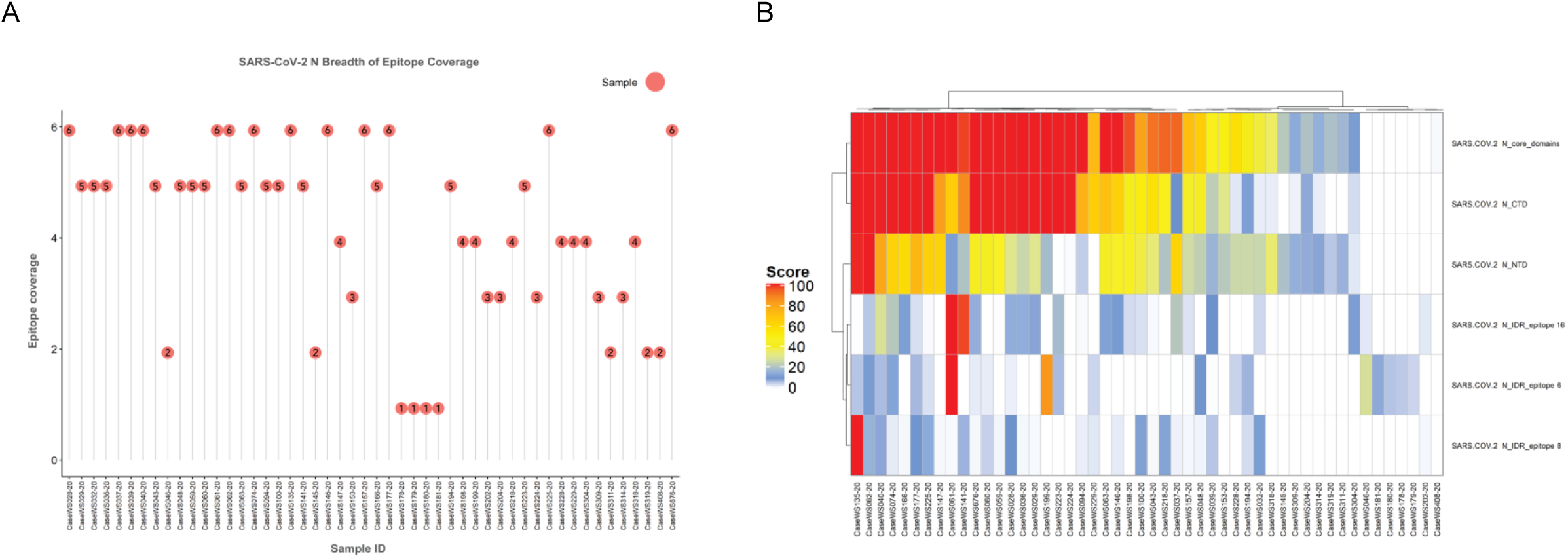
**A-** Overall breadth of epitope coverage in all 50 cases. All positive samples showed antibody reactivity towards at least 1 domain and displayed diverse locality distribution. The numerical value in each red dot represents the number of total antigens with positive antibody reactivity. **B-** Antibody response in COVID-19 positive samples was predominantly directed towards structural domains (N core, NTD and CTD) of the SARS-CoV-2 N protein. The heatmap was plotted based on the ImmuSAFE score. The colour gradient represents the magnitude of score ranging from 0-100.

Eight of the 50 COVID-19 patients exhibited reactivity towards only a single epitope, whereas the remaining 42 COVID-19 samples had reactivity towards multiple epitopes. Figure 2B shows a heatmap representing the diverse antibody profiles of each case sample, with antibody titres predominantly elevated against the folded N core domain, NTD and CTD. The results observed for the 3 IDR epitopes suggest patient-specific immunogenicity with different magnitudes of antibody titre. The magnitude of antibody titres against different regions of the viral N protein may correlate with disease severity and symptom manifestation; this will be reported elsewhere. Interestingly, the present study also found a positive correlation between the age of patients and the magnitude and breadth (the Epitope Coverage score represents the number of N protein domains with positive immunoreactivity) of antibody responses against SARS-CoV-2 N antigens as shown in Figure 3 revealing a significantly broader antibody coverage (Figure 3A) and average reciprocal IgG titre towards all 6 N antigens (Figure 3B) in patients >50 years of age (mean EPC = 5.1), relative to patients aged 20-40 years old (mean EPC = 3). This observation will be further validated in a larger cohort in order to understand its significance in terms of the natural history of COVID-19 disease in different individuals.

**Figure 3:**
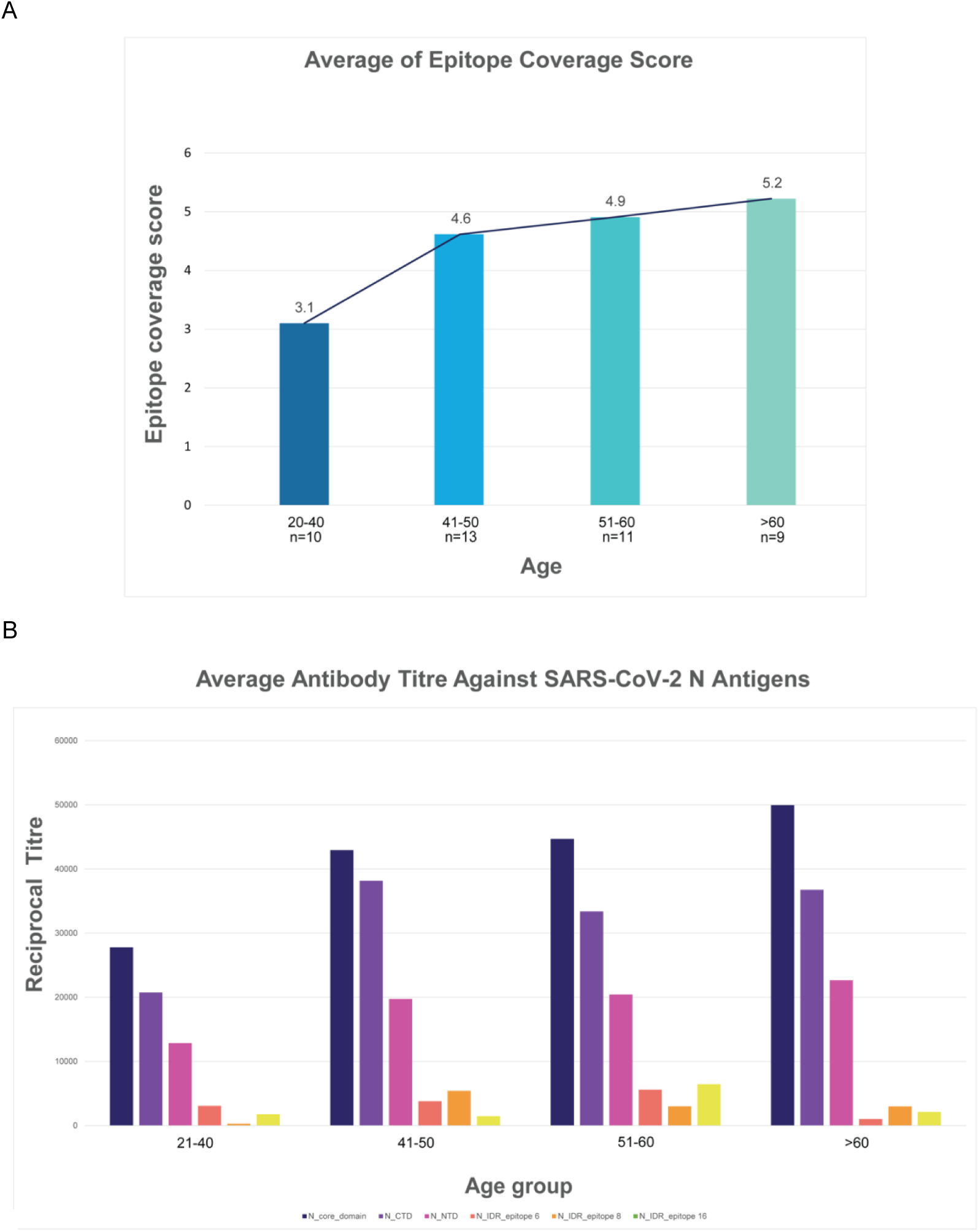
**A-** A positive correlation between increasing age of COVID-19 patients and higher Epitope Coverage scores (EPC). was observed. The figure on the bar plot represents the average number of antigens with positive immunoreactivity exhibited by each age group. **B-** A positive correlation was evident between COVID-19 patients aged above 40 and antibody titre towards SARS-CoV-2 N antigens.

In conclusion, the quantitative ImmuSAFE COVID+ test exhibited 100% specificity and 100 % sensitivity in this study. The inclusion of 6 validated functional N domains of SARS-CoV-2 on our platform ensures that the diversity of antibody reactivity found in different individuals (and the population in general) towards multiple domains of SARS-CoV-2 is captured, leading to increased sensitivity coupled with perfect specificity, whilst also reporting on the localisation and breadth of reactivity between viral antigens and host antibodies. By allowing simultaneous determination of the titres as well as the number and location of epitope targets of anti-SARS-CoV-2 antibodies in individuals, we suggest that our novel, multiplexed, multi-antigen array-based immunoassay platform may thus enable identification of readily-measurable correlates of current high-titre neutralising antibodies, as well as of the likelihood of developing a memory B-cell response that will provide more durable protection against future SARS-CoV-2 infection. Further work to explore this possibility is underway.

## Data Availability

The data that support the findings of this study are available from the Institute of Medical Research (IMR), Ministry of Health, Malaysia, but restrictions apply to the availability of these data, which were used under license for the current study, and so are not publicly available. Data are however available from the authors upon reasonable request and with permission of Institute for Medical Research, Ministry of Health, Malaysia.

## Acknowledgements

We would like to thank the Director General of Health Malaysia for his permission to publish this article. We acknowledge RN Mamat for proof reading of the manuscript, Sengenics Bioinformaticians, KH Azizs, NH Ismail and P Morris for preparation and design of figures.

## Authors contributions

JMB and AA conceived the study. RMZ designed the experiment and provided the samples. NDA and JMB wrote the manuscript. JMB, NDA and TTM designed the content of the array. AJMN cloned and expressed the proteins. MS developed prototype arrays and assays. NSMR, NHB, TNAR, KE and NISB performed the antibody assay. TTM performed the bioinformatics analysis. JMB, AA, RMZ, TTM and NDA interpreted the data. AA and JMB edited the final version of the manuscript.

## Results

Table for main text

Figures for main text

## Supplementary Materials

### Methodology

#### Bioinformatics analysis

##### Image Analysis: Raw Data Extraction

The aim of image analysis is to evaluate the amount of autoantibody present in the serum sample by measuring the median intensities of all the pixels within each probed spot. A raw .tiff format image file was generated for each slide. Automatic extraction and quantification of each spot on the array was performed using the GenePix Pro 7 software (Molecular Devices) which outputs the statistics for each probed spot on the array. This includes the mean and median values of the pixel intensities within a spot and its local background. A GAL (GenePix Array List) file for the array was generated to aid with image analysis. This file contains the information of all probed spots and their positions on the array. Following data extraction, a GenePix Results (.GPR) file was generated for each slide which contains the information for each spot; Protein ID, protein name and foreground and background intensities.

##### Determination of reciprocal titre from fluorescent value output

Data pre-processing and reciprocal titre calculation was performed using the Sengenics ImmuSAFE Cloud-based analysis platform. To perform the analysis, the filename, block number, Name, ID/Protein, foreground median, background median and background standard deviation was consolidated into a template data sheet and cross referenced with identifiers in the sample manifest data sheet. Array replica spots were filtered based on a background plus 2 standard deviation threshold to remove background spots. The mean values for each antigen probe per sample were then calculated. The mean intensities were converted to reciprocal titre values based on the linearity assumption. Reciprocal titre is defined as the reciprocal of the lowest serum dilution that gives signal in the assay. Using a median background signal of 50 RFU as reference, the reciprocal titre for each antigen per sample was calculated and converted into the ImmuSAFE score by rescaling the value to a range between 0-100.

## Notes

### Competing Interest Statement

The authors have declared no competing interest.

### Funding Statement

This study is funded by Malaysia Automotive, Robotics & IoT Institute (MARii), an agency under the Ministry of International Trade and Industry (MITI).

### Author Declarations

Institute for Medical Research, Ministry of Health, Malaysia.

